# Peto odds ratios demonstrate no advantage over classic odds ratios in meta-analysis of binary rare outcomes

**DOI:** 10.1101/2020.10.13.20212290

**Authors:** Chang Xu, Luis Furuya-Kanamori, Lifeng Lin, Suhail A. Doi

## Abstract

In this study, we examined the discrepancy between large studies and small studies in meta-analyses of rare event outcomes and the impact of Peto versus the classic odds ratios (ORs) through empirical data from the Cochrane Database of Systematic Reviews that collected from January 2003 to May 2018. Meta-analyses of binary outcomes with rare events (event rate ≤5%), with at least 5 studies, and with at least one large study (N≥1000) were extracted. The Peto and classic ORs were used as the effect sizes in the meta-analyses, and the magnitude and direction of the ORs of the meta-analyses of large studies versus small studies were compared. The p-values of the meta-analyses of small studies were examined to assess if the Peto and the classic OR methods gave similar results. Totally, 214 meta-analyses were included. Over the total 214 pairs of pooled ORs of large studies versus pooled small studies, 66 (30.84%) had a discordant direction (kappa=0.33) when measured by Peto OR and 69 (32.24%) had a discordant direction (kappa=0.22) when measured by classic OR. The Peto ORs resulted in smaller p-values compared to classic ORs in a substantial (83.18%) number of cases. In conclusion, there is considerable discrepancy between large studies and small studies among the results of meta-analyses of sparse data. The use of Peto odds ratios does not improve this situation and is not recommended as it may result in less conservative error estimation.

## Introduction

Current healthcare decision making relies heavily on evidence from systematic reviews and meta-analyses [1-3]. Meta-analysis pools findings from related studies and provides comprehensive evidence towards answering clinical questions. In meta-analysis, effect estimates of each study are assigned a certain weight and these weighted estimates are then pooled together into a “weighted average effect” [4-6]. Mostly such weights are based on random error (e.g., using the inverse of the study variance), and thus more weights are assigned to larger studies than smaller ones since larger studies have less random error [7, 8]. As a result, larger studies have predominant influence on the pooled effect of a meta-analysis.

Previous studies have investigated the influence of large trials on meta-analysis. Villar et al compared 35 mega-trials to meta-analyses of small studies with the same topics and observed a 20% discrepancy on the direction of the effect size [9]; LeLorier et al compared 12 mega-trials to 19 meta-analyses (there was more than one meta-analysis within the same topic) of small studies and found that in 32.5% of the cases, the directions of the effect sizes were different [10]. This indicates that when the results of mega-trials are added to a meta-analysis of small studies, up to a third of the time, the direction of the conclusion changes - an intervention initially protective, can become harmful (or vice versa); results of smaller studies therefore are discordant with mega-trials in up to a third of cases and may therefore not be robust or useful for clinical decision making.

For meta-analysis of rare events, the discordance between large studies (of the size of a mega-trial) and small studies has not been studied. Meta-analyses of rare events are generally utilized to assess the adverse effects of interventions because single trials have limited power to capture a sufficient number of events [11, 12]. Due to the extremely low frequency of events (even no event), small studies may have larger random or systematic errors in a meta-analysis of rare events and thus their results may be expected to deviate more from that of a large study [13-16]. Currently it is unclear to what extent a large study (or meta-analysis of large studies) may disagree with the results of meta-analysis of small studies for rare event outcomes. We also do not know if the method of computing binary effect sizes can influence this discrepancy. This is because it has been thought that corrections for zero cell counts are not necessary when using Peto’s odds ratios and so it would work well when events are very rare [17].

In this study, we used meta-analyses from the Cochrane Database of Systematic Reviews to empirically explore the extent of agreement between a large study (or pooled large studies if more than one) and the meta-analysis of small studies for rare event outcomes. We also compared meta-analysis results using the Peto odds ratios (no continuity correction) to the classic odds ratios with a continuity correction [15] as many smaller studies will be expected to have study arms with no event.

## Methods

### Cochrane data

The Cochrane Collaboration (https://www.cochrane.org/) is a non-commercial academic organization that aims to provide reliable evidence for health decision. The Cochrane Database of Systematic Reviews (CDSR) contains a comprehensive collection of systematic reviews and meta-analyses, and it has been used for assessing many important meta-analysis issues, including publication bias, missing data and heterogeneity [18-21]. Data from the CDSR were collected from January 2003 to May 2018 using the R package “RCurl” [22].

### Inclusion criteria

Meta-analyses with rare events (binary outcome), with at least 5 studies, and with at least one large study were included. Rare events were defined as incidence rates ≤5% in the control group [23, 24]. Studies with sample size ≥1000 were deemed large; this was akin to the size designated for a mega-trial [10, 25]. To ensure an adequate balance between large and small studies, we excluded the meta-analyses if they had more large studies than small studies. Also, to ensure an adequate influence of larger studies, we excluded the meta-analyses in which the total sample size of large studies was less than that of the smaller studies. Meta-analyses that contained studies with both arms containing zero events were also excluded.

### Data analysis

The Peto and classic ORs and standard errors (SEs) were estimated for meta-analysis of large studies as well as meta-analysis of small studies (excluding large studies). The Peto OR was based on the Yusuf-Peto method [26], and it is recommended by the Cochrane Handbook for meta-analyses of rare events [27]. The Yusuf-Peto method uses the exponent of the difference of observed events to expected events to obtain the asymptotic estimation of the OR; it has been suggested as a valid solution for zero events in one arm [17, 26, 28]. The Peto OR is computed as follows. Consider a 2-by-2 table structure for the data with elements of each cell being *a, b, c, d*, where represent the number of events and non-events in treatment and control groups, respectively. Then the Peto OR (on a log scale, lnOR) and its SE are given by:

- *Peto LnOR* = *{(a* − *E[a]) / V}*
- 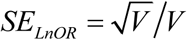
- *V* = *{n*_1_ *n*_2_ *(a* + *c)(b* + *d)} / {N* ^2^*(N* − *1)}*
- *E[a]* = *n*_1_*(a* + *c) / N*
- *n*_1_ = *a* + *b; n*_2_ = *c* + *d; N* = *n*_1_ + *n*_2_

On the other hand, we used the classic OR as an alternative; it applied a continuity correction (cc) of 0.5 for studies with zero event in one treatment arm [15]. It is computed as follows:

- *OR* = *ad / bc*
- 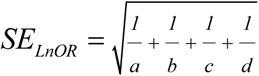
- *OR_cc*= *(a*+ *0*.*5)(d* + *0*.*5)/ (b* + *0*.*5)(c* + *0*.*5)*
- 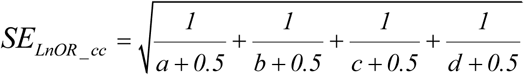

After study ORs were computed, the meta-analysis was carried out using the inverse variance heterogeneity method [7] (IVhet). This method produces better error estimation and is more robust to “small study effects”, compared to the conventional random-effects models, when there is heterogeneity [29]. Also, the normal approximation used by the conventional random-effects model may be poor for small studies or rare events, and this can result in inaccurate estimates [24]. Of note, because of potential heterogeneity, we did not pool studies via the classic fixed-effect approach as is usually done with the Peto approach.

The magnitude and direction of ORs of meta-analyses of large studies versus small studies were compared for each method. The Bland-Altman plot was used to examine the degree of agreement in magnitudes of effect between meta-analysis of small and large studies. This method plots the difference against the average of pooled small and large study results for the lnORs paired by meta-analysis [30]. For the degree of agreement on direction, the kappa statistic was used. The agreement with a kappa within 0-0.4 was treated as weak, 0.41-0.8 as moderate, and 0.81-1.00 as strong [31]. We compared the p-value of meta-analysis of small studies to assess if the two OR methods gave similar results. All data were analysed in Stata/SE 14.0 (Stata, College Station, TX) and the meta-analyses were conducted using the *admetan* command. The statistical significance was pre-specified as 0.05.

## Results

### Data characteristics

From the 61,091 meta-analyses of binary outcome, 52,992 failed to meet the definition of rare events. Of the remaining, 272 had studies with zero events in both arms, 6,879 had less than 5 studies, and 460 did not contain large studies. There were 488 meta-analyses of rare events with large studies. After excluding meta-analyses where the total sample size of large studies was less than the total sample size of smaller studies and excluding meta-analyses with more large studies than small studies, 214 were included for the final analysis (Figure 1).

**Figure 1.**
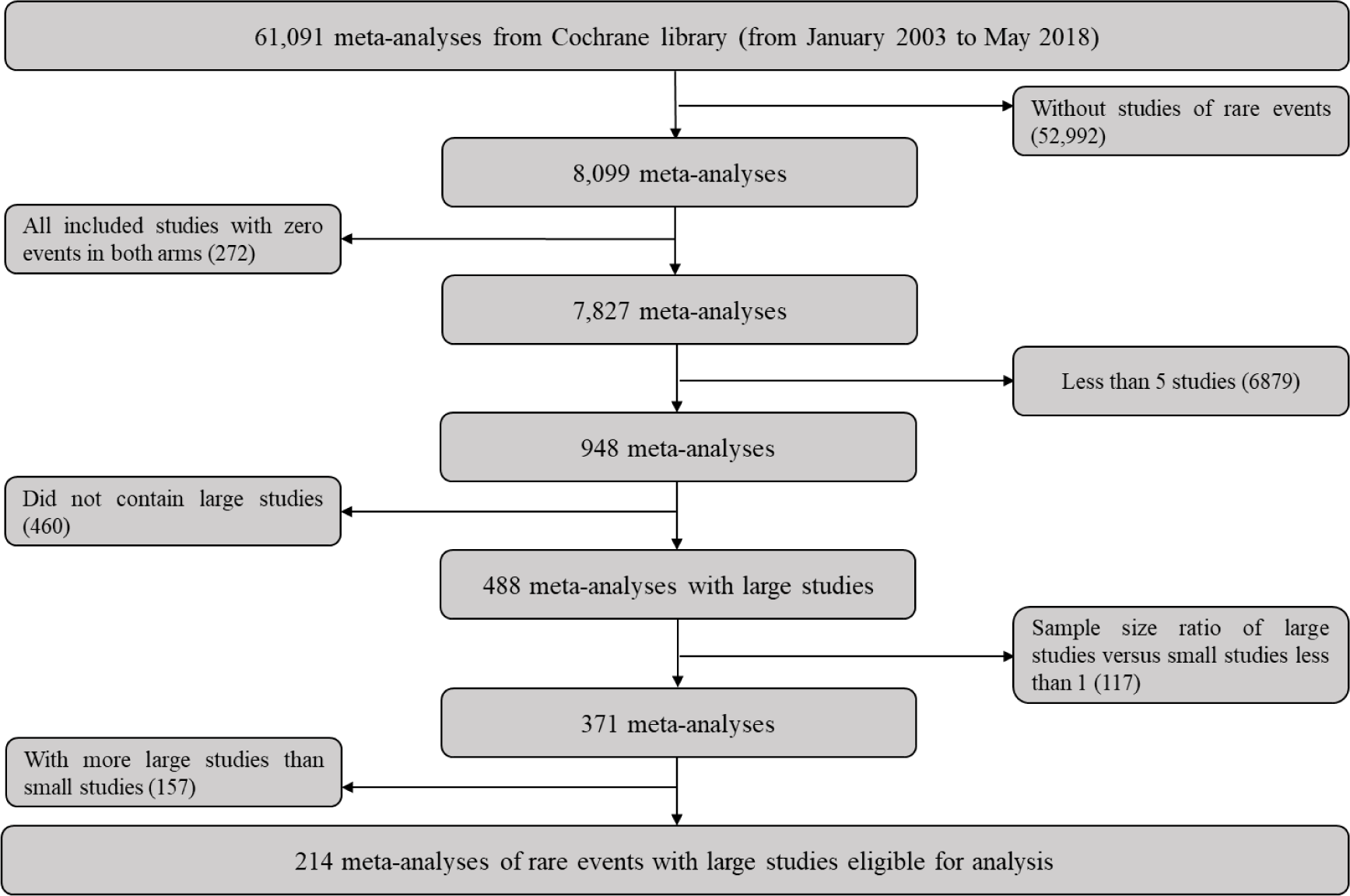
Flow chart of selecting meta-analyses with rare events and large studies.

The number of studies in the meta-analyses ranged from 5 to 18 with a median of 6 (interquartile range [IQR] 5 - 7). The incidence rate in the control group ranged from 0% to 4.92%, with a mean and median of 1.29% (SD 1.24%) and 0.96% (IQR 0.27 - 1.98%), respectively. The sample sizes of small studies ranged from 3 to 730 with a median of 164 (IQR 78 - 299) in the intervention group and from 4 to 498 with a median of 139 (IQR 73 - 258) in the control group. The sample size of the large studies ranged from 53 to 50,053, with a median of 1015 (IQR 678 – 1,678) in the intervention group; and from 82 to 21,555,505, with a median of 952 (IQR 656 –1,706) in the control group. Of note, although individual large study arms had less than 1000 participants, the study size was always ≥1000.

### Comparison of large and small studies

Based on the Peto OR, for meta-analyses of small studies, 121 had an OR > 1 and 93 had an OR <1; whereas for the large studies, 115 had an OR > 1 and 99 had an OR <1. Over the total 214 pairs of ORs (pooled small studies versus pooled large studies), 148 (69.16%) had a concordant direction, while 66 (30.84%) pairs had a discordant direction. The degree of agreement was weak (kappa=0.33, Table 1). The mean difference of the paired lnORs of large studies versus small studies was −0.036 with 12 (5.61%) being outside the limits of agreements (Figure 2).

**Table 1.** Test of agreement on direction for odds ratios for small studies against large studies.

| Peto odds ratios |  |  |  |
| --- | --- | --- | --- |
| Odds ratio (OR) | Large studies |  |  |
| Small studies | (OR < 1) | (OR > 1) | Disagreement rate |
| (OR < 1) | 85 | 36 | 30.84% |
| (OR > 1) | 30 | 63 |  |
| Kappa statistic = 0.22 |  |  |  |
| Classic odds ratios |  |  |  |
| Odds ratio (OR) | Large studies |  |  |
| Small studies | (OR < 1) | (OR > 1) | Disagreement rate |
| (OR < 1) | 82 | 35 | 32.24% |
| (OR > 1) | 34 | 63 |  |
| Kappa statistic = 0.22 |  |  |  |

**Figure 2.**
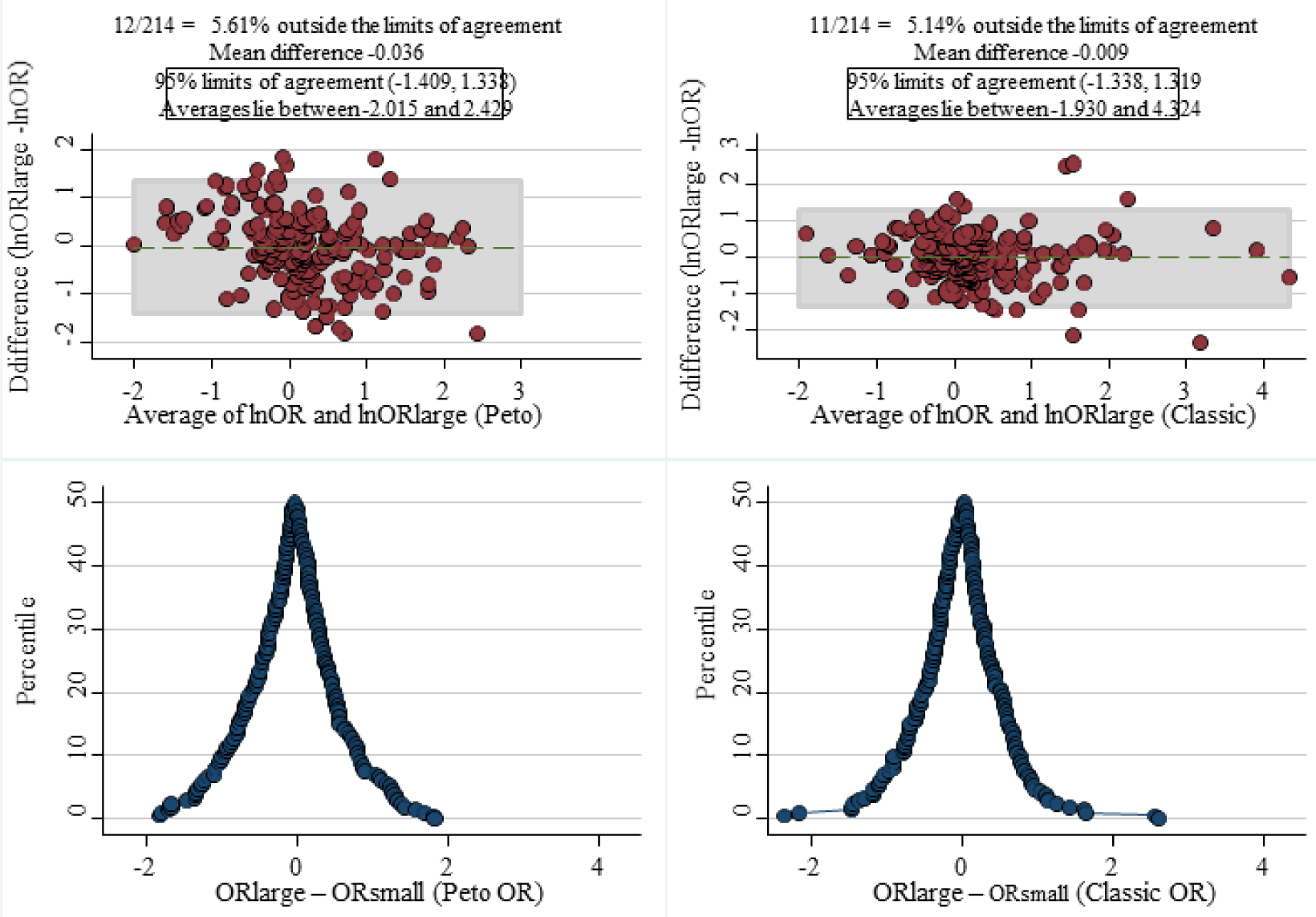
The Bland-Altman (top) and mountain (bottom) plots of the agreement on the magnitude of lnORs. Left: based on Peto method; Right: based on classic method.

Based on the classic OR, for pooled small studies, there were 117 and 97 meta-analyses that had an OR >1 and <1 respectively; and for large studies, 116 and 98 had an OR >1 and <1 respectively. Over the total 214 pairs of ORs, 145 had a concordant direction (67.76%), while 63 pairs had a discordant direction of effect (32.24%). The degree of agreement on direction again was weak (kappa=0.22, Table 1). The mean difference of the paired lnORs of large studies versus small studies was −0.009, with 11 (5.14%) being outside the limits of agreement (Figure 2).

### Comparison of the Peto and classic odds ratios

There was a high consistency on the direction of ORs between the Peto and classic methods. For meta-analyses of large studies, 213 had the same direction and only 1 had different directions (kappa=0.96); for meta-analyses of small studies, only 8 had different directions (kappa=0.96). For the direction of p-values for meta-analysis of large studies, a higher proportion demonstrated statistical significance using the Peto method (65/214 = 30.37%) than using the classic method (55/214 = 25.70%). For small studies, similar results were observed with a higher proportion of significant Peto ORs (55/214 = 25.70%) compared with significant classic ORs (37/214 = 17.29%). Again, there was a strong agreement on the direction of p-values of the Peto and classic methods in both meta-analyses of large studies and small studies (Table 2).

**Table 2.** Test of agreement on direction for odds ratios and p-values between the Peto and classic odds ratios.

| Odds ratios |  |  |  |
| --- | --- | --- | --- |
| Large studies | Peto method |  |  |
| Classic method | (OR < 1) | (OR > 1) | Agreement rate |
| (OR < 1) | 98 | 1 | 99.53% |
| (OR > 1) | 0 | 115 |  |
| Kappa statistic = 0.96 |  |  |  |
| Small studies | Peto method |  |  |
| Classic method | (OR < 1) | (OR > 1) | Agreement rate |
| (OR < 1) | 91 | 6 | 96.26% |
| (OR > 1) | 2 | 115 |  |
| Kappa statistic = 0.91 |  |  |  |
| p-values |  |  |  |
| Large studies | Peto method |  |  |
| Classic method | Positive (p < 0.05) | Negative (p > 0.05) | Agreement rate |
| Positive (p < 0.05) | 55 | 0 | 95.79% |
| Negative (OR > 0.05) | 9 | 150 |  |
| Kappa statistic = 0.93 |  |  |  |
| Small studies | Peto method |  |  |
| Classic method | Positive (p < 0.05) | Negative (p > 0.05) | Agreement rate |
| Positive (p < 0.05) | 37 | 0 | 91.59% |
| Negative (p > 0.05) | 18 | 159 |  |
| Kappa statistic = 0.89 |  |  |  |

### Difference of p-values (meta-analysis of small studies)

The deviation of p-values for pooled small studies (Peto – classic) is presented in Figure 3. In total, the difference in p-values suggested that Peto ORs resulted in spuriously smaller p-values (mean difference = −0.06) compared to classic ORs in a substantial number of cases (83.18%). The deviations from the classic OR p-values were small and only resulted in a cross-over from >0.05 to <0.05 in 18/214 (8.41%) meta-analyses.

**Figure 3.**
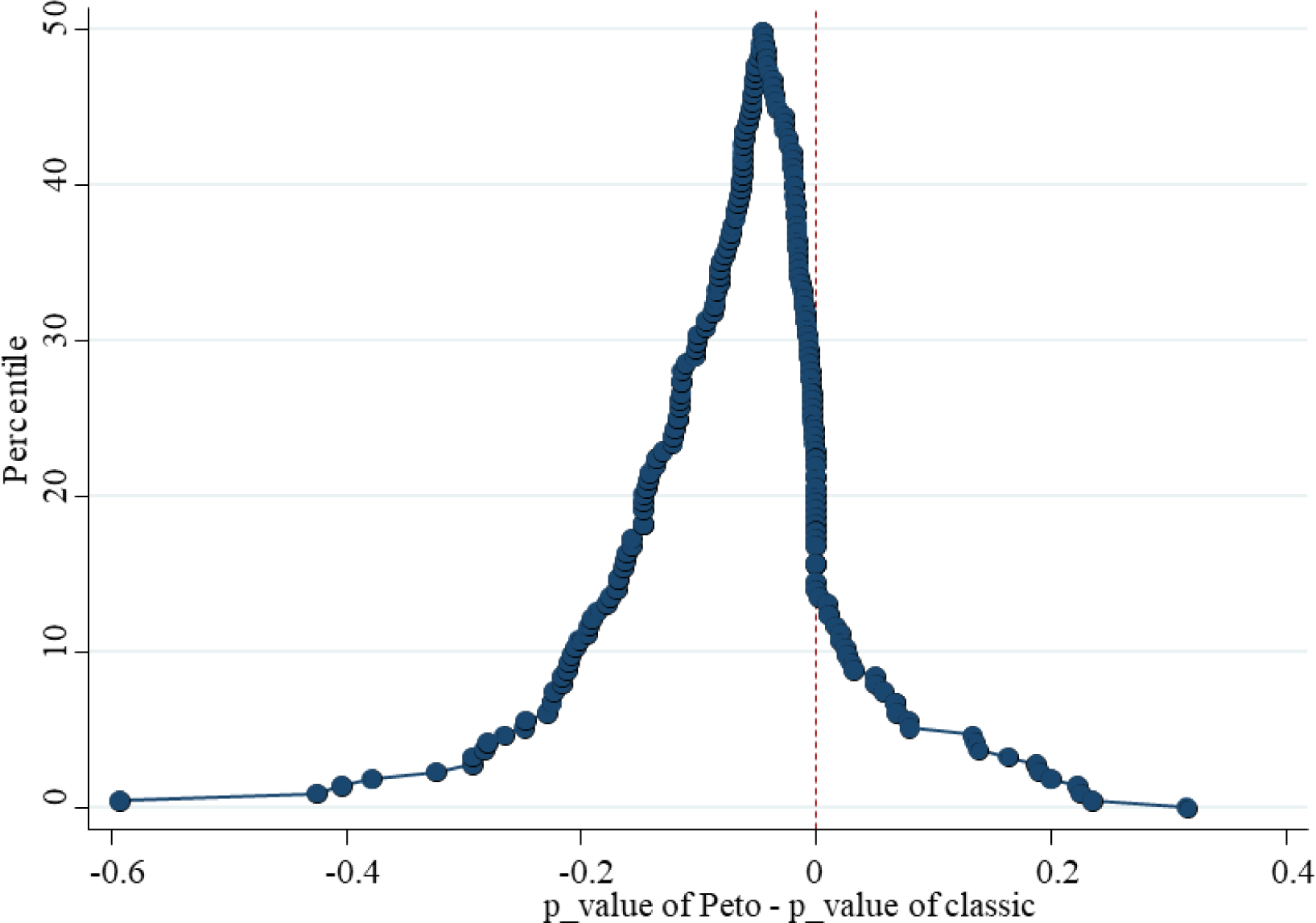
The deviation of p-values for small studies (Peto minus classic). The vast majority (83.18%) of results via Peto demonstrate a spuriously smaller p-value and thus overconfident results. However, only 8.4% of results have a difference sufficient to cross from >0.05 to <0.05.

## Discussion

In this study, we evaluated the extent of agreement and discrepancy of large studies (mega-trial sized) and meta-analysis of small studies for rare event outcomes using empirical data from the CDSR. We found that there was poor agreement in terms of the direction of ORs between large and small trials-about one third of the meta-analyses had a discordant direction of effect. The discrepancy of the effect size between large studies and small studies was similar with the Peto and classic methods of computing ORs. In addition, the differences between the ORs of large versus small trials were comparable across both methods. Where there was more discrepancy in error estimation – the Peto ORs consistently generated smaller p-values and thus had a tendency to produce overconfident results (increase false positive error).

The Peto OR was first proposed as a way to deal with one-arm-zero-events studies by Peto and colleagues in 1985 [26]. An advantage of the Peto OR for such studies is that it does not need a continuity correction to estimate the relative effect and variance. Several simulation studies have shown it to be a more robust effect estimator than the classic OR with continuity correction when the events are rare [15, 28, 32]. We, however, demonstrated no advantage for the Peto ORs in the situation of rare events with large studies; indeed it may lead to less conservative error estimation. The reason our results differ from the existing simulations is that we used real-world data and separated small studies from large studies. Also, we used a more advanced meta-analysis approach (IVhet) that accounts for heterogeneity, rather than the conventional (but unrealistic) fixed-effect model that is associated with the Peto ORs.

For rare events, small studies generally have limited power and insufficient information (zero events) to make a reasonable judgement, even after pooling them together. Large studies tend to have more events allowing a more reliable statistical inference, so some researchers suggest that large studies may be more reliable [33]. This is because meta-analysis incorporates the biases of individual small studies and thus may bring in new sources of bias (e.g. publication bias) [34]. Our study found that one third of smaller studies in a meta-analysis, gave discordant results with the large studies when events are rare. There is therefore a substantial chance of drawing a totally inverse conclusion from small studies when compared to the large studies. This suggests that results of meta-analyses of small studies with sparse results must be viewed with caution.

When there are only smaller studies data available, some additional analyses, such as assessment of publication bias, subgroup analysis and sensitivity analysis, could be potential indicators of the robustness of the results and could provide some guidance. For example, in the case where pooled small studies suggest a positive result, we may need to assess all these other potential sources of uncertainty (additional analyses) before reporting results [35-38]. We can also run a more robust method (e.g. IVhet [7]) as this may be more appropriate to pool the evidence for decision making given the limitations of the random effects method with rare data [24].

### Strengths and limitations

To the best of our knowledge, this is the first empirical study comparing the discrepancy between large studies versus meta-analyses of small studies of rare events. In this study, we used the whole dataset of Cochrane meta-analysis of rare events that also included large studies so our findings are expected to be broadly representative. A strength of this analysis was that we did not use the most popular random-effects meta-analysis estimator given its faulty error estimation [24, 39].

Nevertheless, there are some limitations in the current study. We excluded meta-analyses with studies that contained zero events in both arms. In standard meta-analysis, such studies are also generally excluded; more complicated statistical approaches are needed to properly incorporate the information from such studies in a meta-analysis. In addition, the definition of large trials is somewhat arbitrary. Although this definition has been widely accepted and employed, there was no reasonable theoretical or empirical evidence to support our criterion of N≥1000 for defining large studies. We therefore used additional criteria to define the extent of influence of large studies in this analysis

## Conclusion

There is considerable discrepancy between the results of large studies and small studies with a third of smaller studies disagreeing with large studies. Results of meta-analyses of small studies with sparse results must be viewed with caution. The Peto OR does not improve this situation and may even worsen error estimation and therefore should no longer be considered the method of choice for the meta-analysis of binary data with rare outcomes.

## Data Availability

Data can be obtained from the corresponding authors

## List of abbreviations

OR: Odds ratio
IQR: Interquartile range
CDSR: Cochrane Database of Systematic Reviews
SE: Standard error

## Declarations

### Ethics approval and consent to participate

Not applicable

### Consent for publication

All authors approved for publication

### Data sharing

Data can be obtained by contacting the corresponding author. The authors downloaded the data from Cochrane Database of Systematic Reviews (CDSR) under the full access through Florida State University (in 2018). Please verify the accessibility on CDSR of your institution in order to get the data.

## Acknowledgements

None

## Authors’ contributions

SD and XC conceived and designed the study; LL conducted the data collection; XC, LFK, and SD programmed the code for data cleaning and analyzed the data; XC drafted the manuscript; SD and LFK provided statistical guidance; SD, LFK and LL provided critical comments, revised the manuscript, and approved the final version.

